# Varicella Zoster infection manifesting as shingles is not independently associated with ischemic nor hemorrhagic stroke using the large Nationwide Inpatient Sample (NIS) database

**DOI:** 10.1101/2025.09.24.25336593

**Authors:** Mehrnoosh Hashemzadeh, Bryce Renzi, Mehrtash Hashemzadeh, Mohammad Reza Movahed

## Abstract

**Background:** Previous studies have shown a correlation between having had a previous infection of Varicella Zoster, and an increase in risk of stroke, even in the absence of rash. Our goal was to further evaluate this correlation using a large, publicly available inpatient database, searching for patients who have had either shingles, a stroke, or both.

**Method:** We used the National (Nationwide) Inpatient Sample (NIS) to assess for any correlation between active shingles and increased stroke risk, looking at patients who were admitted to an inpatient setting in 2016-2020. Uni- and multivariate analysis was performed adjusting for comorbid conditions

**Results:** A total of 4,115,724 patients ≥ 18 years old had a diagnosis of stroke. From them, 2.85% had a history of shingles vs 2.77% without a history of shingles (OR, 1.03; 95% CI, 0.97-1.09; p = 0.31). When comparing only for hemorrhagic stroke, the percentage of patients admitted who had a history of shingles was 0.89%, versus 0.74% of patients without a history of shingles (OR, 1.20; 95% CI, 1.09-1.33; p = <0.001). However, after multivariate analysis, this difference became statistically insignificant (OR: 0.91, CI: 0.85-1,05, p =0.32)

**Conclusion:** **W**e found no correlation between having a history of shingles as increased risk of hemorrhagic of ischemic stroke over the five years we studied.

## Introduction

Varicella Zoster Virus (VZV), the cause of chickenpox and shingles, is a highly infectious double-stranded DNA virus that can spread via respiratory droplets, which can cause headache and fever, as well as complications such as pneumonia and encephalitis. However, what makes it especially notable is its ability to lay dormant after the initial infection within neurons of the trigeminal and dorsal root ganglia, potentially re-activating years later during periods of stress or immunocompromise to cause shingles (1), which is a dermatomal rash which causes severe pain. VZV is also the only known virus that can “replicate in arteries and produce vasculopathy”, (2) and can do so even in the absence of a rash. (3)

A stroke is a potentially fatal event of the brain from either hypoperfusion or from hemorrhage. Even without immediate death, complications can be severe such as loss of bodily functions and senses. Patients who have experienced a stroke have significantly worse functional status and quality of life when compared to those who have not had a stroke. (4) Prophylaxis and secondary prevention of stroke in those at higher risk can decrease morbidity and increase the quality of life for these patients.

Over 99% of people born prior to 1980 in the United States have been at one point infected with Varicella Zoster. Approximately 1 in 3 people in the United States will develop herpes zoster during their lifetime. (5) While there has been a vaccine for varicella since 1995 and a shingles vaccine since 2006, there are still breakthrough cases of both chickenpox and shingles as the vaccines are not 100% effective. The varicella vaccine has an efficacy of 92% (6) and the shingles Shingrix vaccine has an efficacy of 97.2% among those aged 50 and older. (7)

The lifetime risk of having a stroke, either ischemic or hemorrhagic, is 23.8% in the United States. (8) Previous epidemiological studies of different populations have shown an increased risk of stroke for some period of time after having VZV. One cohort study consisting of all adults in Denmark aged 18 and older showed an increased risk of stroke after VZV, with the increased risk decreasing over time down to a %5 increase at the one-year mark. (9) One cohort study of a county population in Minnesota aged 50 and older showed an increased risk of stroke up to 3 months after having shingles when controlling for confounding factors. (10)

The goal of this study was to determine if there was a significant correlation between shingles and stroke in the inpatient setting, with the hope of improving outcomes and prophylactically treating for accurate risk.

## Methods

### Data Collection and Data Sources

As a data source, we used the National (Nationwide) Inpatient Sample (NIS), which is part of a suite of tools developed under the Healthcare Cost and Utilization Project (HCUP). (11) The NIS is an ‘all-payer’ inpatient healthcare database that contains data from 1988 through 2020, and currently stores data from over 7 million hospital visits annually, making it the largest public database of its type. This database is publicly available for analysis, and because it is de-identified data, it is also exempt from IRB review. Despite being de-identified, the database contains a wealth of information on each hospital stay, including ICD-9 or ICD-10 codes depending on the year, length of stay, patient demographics, and comorbidity measures.

### Collected Information

This study included patients in the NIS database from the years 2016 through 2020. This is because 2016 is the first full year of ICD-10 codes, which were used to select patients, and 2020 is currently the last full year of data to which we have access. Only patients 18 and older were included in the study. The ICD-10 code used to select patients for shingles was B029 (Zoster without complications). The ICD-10 codes used to select patients for stroke were 92 different I63 codes (Cerebral infarction).

### Statistical Analysis

Using STATA 17 (Stata Corporation, College Station, TX), retrospective uni- and multivariate analysis was performed on patients who met inclusion criteria between and including 2016 and 2020. Focus was given to any potential correlation between shingles and stroke while considering age, sex, race, and comorbidities such as hypertension, diabetes, hypercholesteremia, and chronic kidney disease. Chi-square testing and Fisher’s Exact Test were used to compare categorical variables, whereas Logistic Regression was used to compare odds of clinical outcomes based on categorical data. All p-values are 2-sided and a p-value ≤ 0.05 was statistically significant. 95% confidence intervals and means were calculated for all relevant analyses.

## Results

A total of 4,115,724 patients with stroke were included in the study, of which 6,420 had a history of shingles, and 4,109,304 had no history of shingles. Aggregating all years measured (2016-2020), the percentage of patients admitted for ischemic or hemorrhagic stroke who have or have had shingles was 2.85%, versus 2.77% of admitted patients having a stroke without a diagnosis of current or past shingles (OR, 1.03; 95% CI, 0.97-1.09; p = 0.31). Broken up by year, 2016 and 2018 showed the largest differences with a p of 0.23 and 0.03 respectively. 2017 showed the largest p-value with p=0.86 (Table 1; Figure 1).

**TABLE 1:**
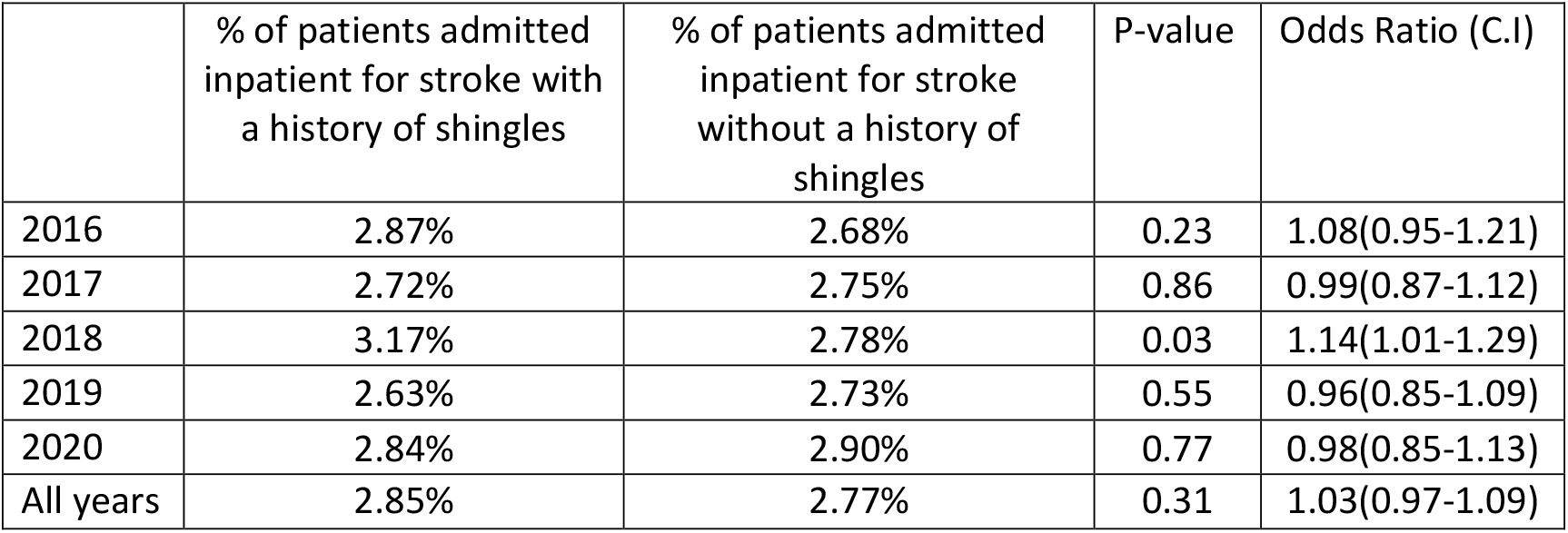
Prevalence of Admission for Stroke among Patients with and without a History of Shingles, 2016-2020.

|  | % of patients admitted<br>inpatient for stroke with<br>a history of shingles | % of patients admitted<br>inpatient for stroke<br>without a history of<br>shingles | P-value | Odds Ratio (C.I) |
| --- | --- | --- | --- | --- |
| 2016 | 2.87% | 2.68% | 0.23 | 1.08(0.95-1.21) |
| 2017 | 2.72% | 2.75% | 0.86 | 0.99(0.87-1.12) |
| 2018 | 3.17% | 2.78% | 0.03 | 1.14(1.01-1.29) |
| 2019 | 2.63% | 2.73% | 0.55 | 0.96(0.85-1.09) |
| 2020 | 2.84% | 2.90% | 0.77 | 0.98(0.85-1.13) |
| All years | 2.85% | 2.77% | 0.31 | 1.03(0.97-1.09) |

**Figure 1:**
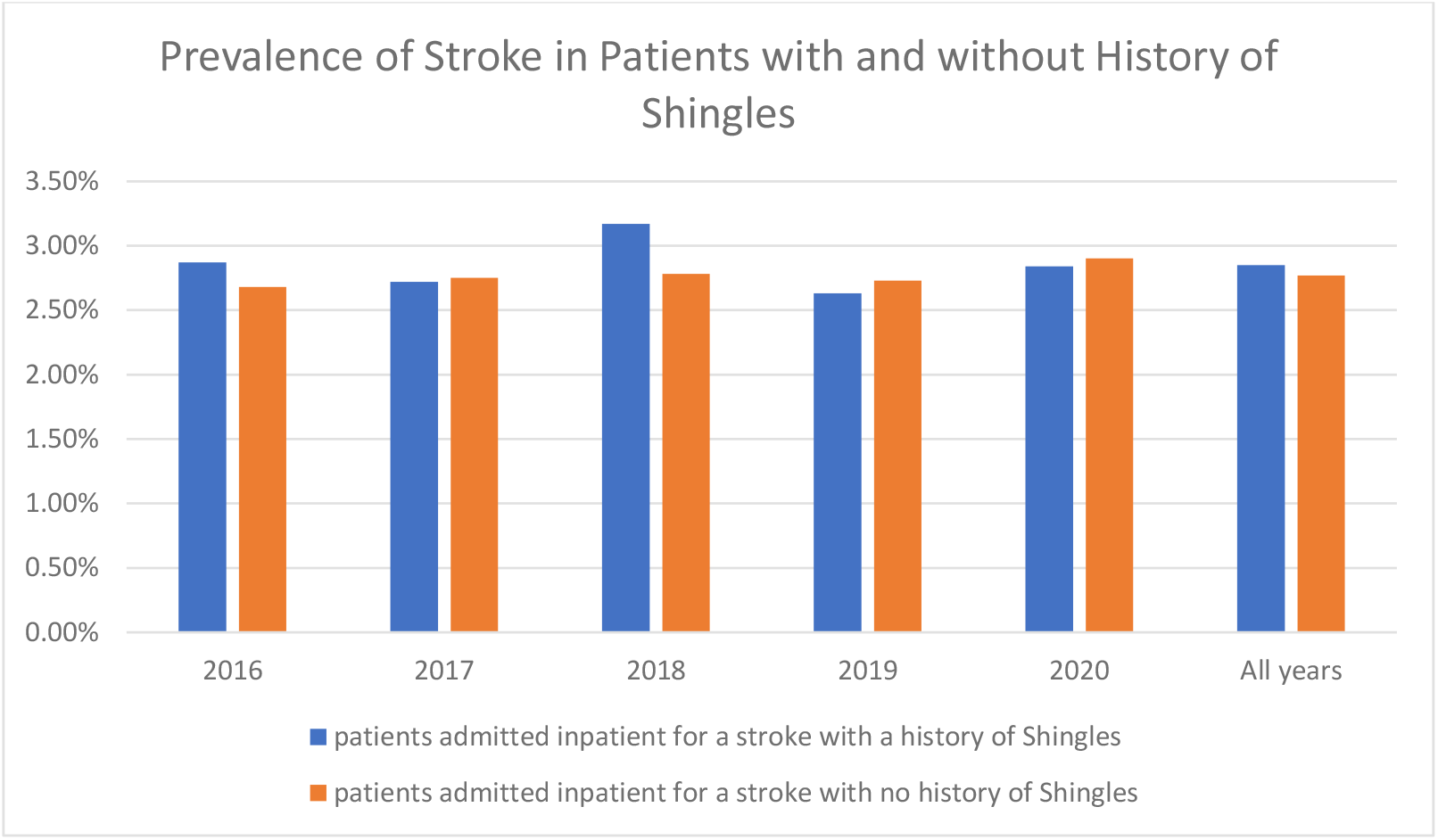
Prevalence of Stroke in Patients with and without History of Shingles, 2016-2020.

When separating data by hemorrhagic vs. ischemic stroke, there is a significant difference in prevalence. For ischemic stroke, there are no years with a significant difference in the percentage of patients with active or past shingles who have a stroke versus the percentage of patients with no history of shingles who have a stroke (p=0.31). For hemorrhagic stroke aggregating all years measured, the percentage of patients admitted with a history of shingles was 0.89% versus 0.74% in those with no history of shingles (p=<0.001) (Table 2; Table 3). However, after adjusting for comorbid conditions in the multivariate analysis, hemorrhagic stroke was not associated with shingles and this difference became statistically insignificant (OR: 0.91, CI: 0.85-1,05, p =0.32)

**Table 2:** Prevalence of hemorrhagic Stroke in Patients with and without History of Shingles, 2016-2020.

|  | % of patients admitted inpatient for hemorrhagic stroke with a history of shingles | % of patients admitted inpatient for hemorrhagic stroke with no history of shingles | P-value | Odds Ratio (C.I) |
| --- | --- | --- | --- | --- |
| 2016 | 0.81% | 0.68% | 0.12 | 1.19(0.96-1.48) |
| 2017 | 0.78% | 0.70% | 0.35 | 1.12(0.88-1.41) |
| 2018 | 1.09% | 0.75% | <0.001 | 1.47(1.20-1.80) |
| 2019 | 0.82% | 0.77% | 0.56 | 1.07(0.85-1.36) |
| 2020 | 0.98% | 0.83% | 0.18 | 1.18(0.93-1.51) |
| All years | 0.89% | 0.74% | <0.001 | 1.20(1.09-1.33) |

**Table 3:** Prevalence of ischemic Stroke in Patients with and without History of Shingles, 2016-2020.

|  | % of patients admitted inpatient for ischemic stroke with a history of shingles | % of patients admitted inpatient for ischemic stroke with no history of shingles | P-value | Odds Ratio (C.I) |
| --- | --- | --- | --- | --- |
| 2016 | 2.12% | 2.10% | 0.91 | 1.01 (0.87-1.16) |
| 2017 | 2.08% | 2.17% | 0.58 | 0.96 (0.84-1.11) |
| 2018 | 2.30% | 2.18% | 0.44 | 1.06 (0.92-1.21) |
| 2019 | 1.91% | 2.11% | 0.18 | 0.90 (0.78-1.05) |
| 2020 | 1.99% | 2.23% | 0.16 | 0.89 (0.76-1.05) |
| All years | 2.08% | 2.16% | 0.31 | 0.97(0.90-1.03) |

There was a statistically significant difference in age between stroke patients with shingles (72.03 SD=13.46) versus controls (69.10 SD=14.54) (p<0.001). There was also a statistically significant (p=<0.001) difference in patient population controlling for gender, with 57.68% of stroke patients with active shingles being female, and 42.32% being male. This is compared to 48.97% of total stroke patients being female, and 51.03% being male. Baseline characteristics can be seen in table 4.

**Table 4:** Average Age, sex, and race of Stroke Patients with and without History of Shingles.

|  | Shingles History | No Shingles History | P-value |
| --- | --- | --- | --- |
| Age (Mean±SD) | 72.03±13.46 | 69.10±14.54 | <0.001 |
| Gender |  |  | <0.001 |
| Male | 42.32% | 51.05% |  |
| Female | 57.68% | 48.95% |  |
| Race |  |  | <0.001 |
| White | 70.08% | 66.90% |  |
| Black | 12.18% | 17.26% |  |
| Hispanic | 10.56% | 8.95% |  |
| Asian/Pac Isl | 4.03% | 3.47% |  |
| Native American | 0.40% | 0.50% |  |
| Others | 2.75% | 2.92% |  |

## Discussion

Stroke is a potentially fatal event, for which there are multiple abortive and preventive pharmacologic and non-pharmacologic regimens.

Multiple mechanisms have been proposed for how VZV could increase the risk of stroke. (12) One possible mechanism is nonspecific inflammation caused by the virus as well as a protein S deficiency related to the virus. One case series postulated that VZV could sometimes stimulate the production of an antibody that affects protein S. (13) An alternative mechanism proposed is that VZV causes “virtually identical pathological changes [] in the arteries” to those seen in patients with giant cell arteritis, and granulomatous changes in “all layers of cerebral arteries”. (3) The virus is able to do this as it can spread from the trigeminal afferent fibers into the brain ‘transaxonally.’ (3) Imaging has shown vessel wall thickening in patients with VZV vasculopathy. In one study, patients with VZV vasculopathy confirmed by antibodies in the CSF had improved imaging findings with the use of acyclovir. (3) One study which studied the pathology of early and late VZV vasculopathy from biopsies showed “T cells expressing CD3 as well as cells expressing CD4, CD8, CD20, and CD68 [as] present in the adventitia associated with a thickened intima in early VZV vasculopathy.” (14) This study also suggested that “neutrophils may play a role”, via production of reactive oxygen species leading to “loss of vascular smooth muscle cells” and production elastase further weakening the vessel wall. (14)

The current literature regarding the effect of shingles on stroke is scarce and has differing inclusion and exclusion criteria as well as controversial outcomes. One meta-analysis showed that “[herpes zoster] was significantly correlated with increased risk of stroke, and the pooled RR was 1.36 (95% confidence interval [CI]: 1.10, 1.67) (P =.004).” (15) In addition, a stronger RR of 4.42 was shown for those with a history of herpes zoster ophthalmicus. (15) Of note this meta-analysis did not show an association for hemorrhagic stroke when analyzed separately. (15) One retrospective case-control study done using a database of Veterans Affairs patients showed an increased risk of stroke within 30 days of an active infection “(odds ratio [OR], 1.93 [95% confidence interval {CI}, 1.57-2.4]; P <.0001)” (16). One factor that may affect results and studies is that infection without rash may still later result in TIA or stroke. (17)

This study was the first to examine data from the National Inpatient Sample (NIS) database, focusing solely on patients who were admitted to the inpatient service of a hospital for their stroke. In addition, patients were categorized based on ICD-10 codes input as primary reason for admission versus any secondary diagnosis that was not the primary reason for admission.

After multivariate analysis, our study found no significant correlation between having an active shingles infection and risk of stroke. Possible reasons for the discrepancy between our results and those of some of the current literature include differing patient populations, as well as different methods of identifying patients who fit each study group. If a patient’s primary care is not within the same health system as where they are receiving acute stroke care, a history of shingles may not be accurately input into the EMR with the appropriate ICD-10 code even if such a history does exist. In addition, our patient population is solely drawn from inpatient admissions, whereas other studies may draw from patients seen in an outpatient setting as well. One possible confounding factor is that if a patient had a stroke and died before being admitted to the hospital, that patient would not be included in our data. Of note, before adjusting for variables such as age, sex, race, and comorbidities, the data showed patients with a history of shingles had a statistically significant increased risk of hemorrhagic stroke as compared to those with no history of shingles.

### Limitations

This retrospective study used data from only a publicly available inpatient database. Since this database is based only on inpatient data, this study may not reflect accurately data from outpatient experiences. Furthermore, since this data was collected from multiple centers, different centers may have different ICD-10 coding practices which may affect results. Our study only focuses on adults aged 18 and older; pediatric data may differ.

## Conclusion

Based on our results, we found no correlation between having a history of shingles and having an increased risk of ischemic or hemorrhagic stroke over the five years studied. Future studies are needed to further elucidate any correlation between Varicella Zoster Infection and stroke.

## Data Availability

NIS Database is publicly available

